# Effect of a Mobile-based Intervention on Mental Health in Frontline Healthcare Workers Against COVID-19: Protocol for a Randomized Controlled Trial

**DOI:** 10.1101/2020.11.03.20225102

**Authors:** Maria J. Serrano-Ripoll, Ignacio Ricci-Cabello, Rafael Jiménez, Rocío Zamanillo-Campos, Aina M. Yañez Juan, Miquel Bennasar-Veny, Carolina Sitges, Elena Gervilla, Alfonso Leiva, Javier García-Campayo, Maria Esther García-Buades, Mauro García-Toro, Guadalupe Pastor Moreno, Isabel Ruiz-Perez, Pablo Alonso-Coello, Joan Llobera Canaves, Maria A. Fiol-Deroque

## Abstract

**Aim:** To evaluate the impact of a psychoeducational, mobile health intervention based on cognitive behavioural therapy and mindfulness-based approaches on the mental health of healthcare workers at the frontline against COVID-19 in Spain.

**Design:** We will carry out a two-week, individually randomised, parallel group, controlled trial. Participants will be individually randomised to receive the PsyCovidApp intervention or control App intervention.

**Methods:** The PsyCovidApp intervention will include five modules: emotional skills, lifestyle behaviour, work stress and burnout, social support, and practical tools. Healthcare workers having attended COVID-19 patients will be randomized to receive the PsyCovidApp intervention (intervention group) or a control App intervention (control group). A total of 440 healthcare workers will be necessary to assure statistical power. Measures will be collected telephonically by a team of psychologists at baseline and immediately after the two weeks intervention period. Measures will include stress, depression and anxiety (DASS-21 questionnaire – primary endpoint), insomnia (ISI), burnout (MBI-HSS), post-traumatic stress disorder (DTS), and self-efficacy (GSE). The study was funded in May 2020, and was ethically approved in June 2020. Trial participants, outcome assessors and data analysts will be blinded to group allocation.

**Discussion:** Despite the increasing use of mobile health interventions to deliver mental health care, this area of research is still on its infancy. This study will help increase the scientific evidence regarding the effectiveness of this type of intervention on this specific population and context.

**Impact:** Despite the lack of solid evidence about their effectiveness, mobile-based health interventions are already being widely implemented because of their low cost and high scalability. The findings from this study will help health services and organizations to make informed decisions in relation to the development and implementation of this type of interventions, allowing them pondering not only their attractive implementability features, but also empirical data about its benefits.

**Clinical trial registration:** NCT04393818 (ClinicalTrials.gov identifier)

## INTRODUCTION

The current COVID-19 pandemic is posing unprecedented challenges for health systems and healthcare workers (HCWs) alike. Worldwide, HCWs are facing increased workloads, are at high risk of infection (for themselves and their cohabitants)^1-3^, and lack of resources to handle the situation. As a result of having to make decisions such as how to provide care for severely unwell patients with constrained or inadequate resources, or how to balance their own physical and mental healthcare needs with those of patients, they are suffering a moral injury^4^. This extreme situation has important implications for HCW’s mental health^5^. A recent systematic review examining the mental health problems among frontline HCWs during viral epidemic outbreaks^5^ observed a high prevalence of acute stress (40%), anxiety (30%), burnout (28%), depression (24%) and post-traumatic stress disorder (13%). Health services worldwide are implementing strategies to mitigate these psychological consequences, most of which are based on the provision of cognitive-behavioural therapy (CBT) (e.g. United States^6 7^, France^8^, Italy^9^, Sierra Leone^10^). However, there is still very limited empirical evidence about the effectiveness of available interventions to protect mental health of HCWs during viral pandemics^5^.

Mobile health (mHealth) interventions are rapidly gaining popularity because of their low cost, high scalability and sustainability features. Recent trials have examined the efficacy of mHealth interventions addressing mental health problems, including suicide^11^, schizophrenia^12^, substance use disorders^13^, and psychosis^14^, among others^15^. Recent systematic reviews investigating the efficacy of standalone smartphone apps for mental health show that, although they have potential for improving mental health symptoms, the available evidence is still scarce and more rigorous trials are needed ^15 16^.

## BACKGROUND

mHealth interventions are well suited to help HCWs to combat the adverse effects of working in such high-pressure situations for a prolonged time period for two reasons^17^. First, they can address non-treatment-seeking behaviour (a common issue among HCWs^18^), as they provide the opportunity to engage individuals in need of treatment timely and anonymously by providing portable and flexible treatment. Second, they are delivered in absence of face-to-face interactions, reducing the risk of infection for SARS-CoV-2. However, their effectiveness in this specific context and population is largely unknown: As observed by a recent review^19^, only 27% of the studies about mental health apps to assist HCW during COVID-19 included empirical evaluation of the reported interventions. Robust, large scale trials are, therefore, urgently needed to determine the extent to which mHealth interventions can improve mental health of frontline HCWs.

Spain is the country with higher mobile phone use rates in the world, with 98% of users^20^. On May 8 of 2020, Spain reported the highest cumulative number of COVID-19 infections among HCWs around the world (30,663 infections - counting for 20% of all HCW infections worldwide)^21^. The pandemic produced very severe consequence in Spanish HCWs’ mental health, with around 57% of HCWs presenting symptoms of posttraumatic stress disorder, 59% of anxiety disorder, 46% depressive disorder, and 41.1% feeling emotionally drained^22^. Under these exceptional circumstances, we received funding to develop and evaluate a CBT and mindfulness-based intervention using an mHealth, to protect mental health of Spanish HCWs attending the COVID-19 emergency.

This article describes the protocol for the PsyCovidApp trial. This protocol trial has been prepared in accordance with the Standard Protocol Items: Recommendations for Interventional Trials (SPIRIT) guidelines^23^.

## THE STUDY

### Aim

The aim of this study is to evaluate the effectiveness of a psychoeducational mHealth intervention against a control App intervention, to protect mental health in HCWs at the frontline against COVID-19 in Spain.

### The specific objectives are

- To analyse the efficacy of the intervention (in the overall sample and in specific subgroups based on baseline mental health status and use baseline use of psychotherapy and psychopharmaceutical drugs) in reducing the levels of depression, anxiety, anxiety, acute and posttraumatic stress, burnout, self-efficacy, and insomnia.
- To examine the usability of the intervention.

### Design

We will carry out a blinded, two-weeks, individually randomised, parallel group, controlled trial. Participants will be individually randomised with an allocation ratio of 1:1 to receive either the PsyCovidApp intervention or control App intervention (both described below).

### Setting

The trial will be carried out in healthcare centres in Spain, including hospitals, primary care centres, and care homes.

### Participants

The trial will include male and female HCWs aged>18, who report having provided healthcare to patients with COVID-19 during the viral outbreak in Spain (from the onset of the health emergency to the recruitment time). For this study HCWs will be defined as professionals regulated by a Health System who deliver care and services whose primary intent is to enhance health. HCWs from any medical speciality (pneumology, internal medicine, emergency, primary care, etc.) and role (doctors, nurses, nurse assistants, etc.) with access to a smartphone will be included. We will include HCWs who have provided direct, face to face, healthcare to patients with a diagnosis of infection by COVID-19. This will include healthcare to any health problem patients may experience (i.e., not only caused by COVID-19). We will exclude HCWs with no access to a smartphone, or not able to download and activate the app used to deliver the intervention during the next 10 days following the baseline assessment in their smartphone.

### Withdrawal criteria

HCWs will be considered withdrawn from the study if they retire their consent to participate, or if they do not receive a postintervention evaluation within the next 15 days after the end of the intervention period. Reasons for withdrawals and discontinuation of any participant from the trial will be recorded.

### Procedure

A flowchart describing the PsyCovidApp trial procedures is available in Figure 1. We will send invitations to HCWs to participate in the trial through social media and key stakeholders hospital managers and communication departments, trade unions of HCWs, scientific societies, research institutes, private insurances companies, home care centres and professional colleges). HCWs willing to participate will register their interest by completing an online questionnaire, which will contain a participant information sheet. A team of psychologists, who will have previously received a 1-hour training session (to ensure homogeneity in recruitment, questionnaire administration, and data entry methods), will contact via telephone with registered HCWs to confirm eligibility criteria, to obtain informed consent (audio-recorded), to carry out a psychological (pre-intervention) evaluation, and to instruct participants about how to download the Clinicovery© App (Apploading, Inc).Clinicovery© is a platform that allows uploading information in multiple formats (text, video, audio) and organize it in modules^24^. These modules are then made available to users of the Clinicovery© App. This system has two main advantages that makes it ideally suited for its use in clinical trials: first, the access to the app contents remains under the control of the researchers, who individually activate the contents of the app after HCWs have registered (i.e., users cannot access to the intervention with no activation from a member of the research team); second, it allows the researchers to allocate different contents to different groups of users (e.g., intervention and control groups).

**Figure 1.**
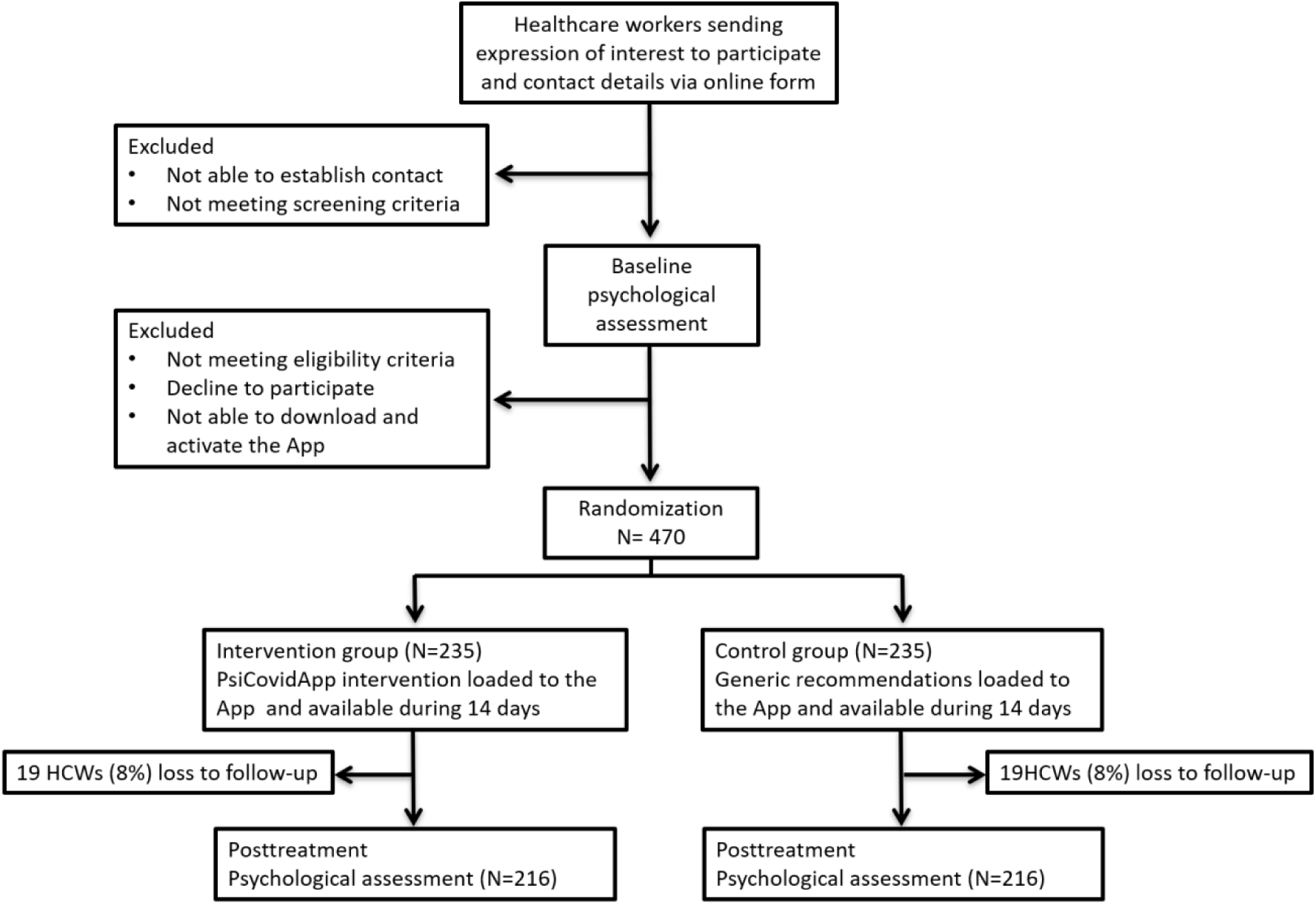
Description of the PsyCovidApp trial.

Within 48 hours after participants successfully download and activate the App (user activation of the App will be used as a checkpoint to ensure participants can successfully use it), a member of our research team will load the contents to the App according to the group participants have been allocated to. This procedure will ensure allocation concealment. During the next 14 days, all HCWs will continue with their usual care (e.g,. use of psychopharmaceutical drugs or psychotherapy, if any) throughout the study. In addition, the intervention group will have access to contents of the PsyCovidApp intervention (described below). Participants in the control group will only have access to control App intervention (below). After two weeks, the contents uploaded in both groups will be disabled, and a post-intervention psychological assessment will be undertaken via telephone. After the post-intervention assessment, all participants will be offered free, unrestricted access to the PsyCovidApp intervention.

### Randomization and blinding

We will randomize patients individually using a computer-generated sequence of random numbers. HCWs will be blinded to group allocation. Data analysts and outcome assessors (in this case, the psychologists who will undertake the pre- and post-intervention psychological evaluations) will also be blinded.

### Description of the PsyCovidApp intervention

The PsyCovidApp intervention was developed on May 2020 by a group of nine experts (five psychologists, two psychiatrists, and two experts in lifestyle modification), informed by findings from an exploratory qualitative study involving in-depth interviews with eight HCWs seeking psychological support, as a result of their professional activity during the COVID-19 pandemic (unpublished results). The intervention developers adhered to current recommendations for the development of mental health Apps^25^. The PsyCovidApp intervention aims to prevent and mitigate the most frequent mental problems suffered by HCWs attending the current COVID-19 emergency (depression, anxiety, stress, and burnout). The intervention includes psychoeducational components, and it is based on CBT and mindfulness approaches. The contents are grouped into five main sections (see Box 1): emotional skills, lifestyle behavior, work stress and burnout, social support, and practical tools. Each section contains multiple modules, covering the following areas: i) monitoring mental health status; ii) educational materials about psychological symptoms (e.g. anxiety, worry, irritability, mood, stress, moral distress, etc.); iii) practical tips to manage pandemic-related stressors (e.g., mindfulness, relaxation and breathing techniques, coping strategies, survival skills to emotional crises); iv) healthy lifestyles and practical tips to promote them; v) organizational and individual strategies to promote resilience and reduce stress at work and the burnout syndrome, and; v) promotion of social support. The contents are displayed using written information, audios and videos (see Figure 2). Additional information is offered through links to web pages, articles, guides, videos and audios. All these contents will be permanently available during the 2-week intervention period.

**Figure 2.**
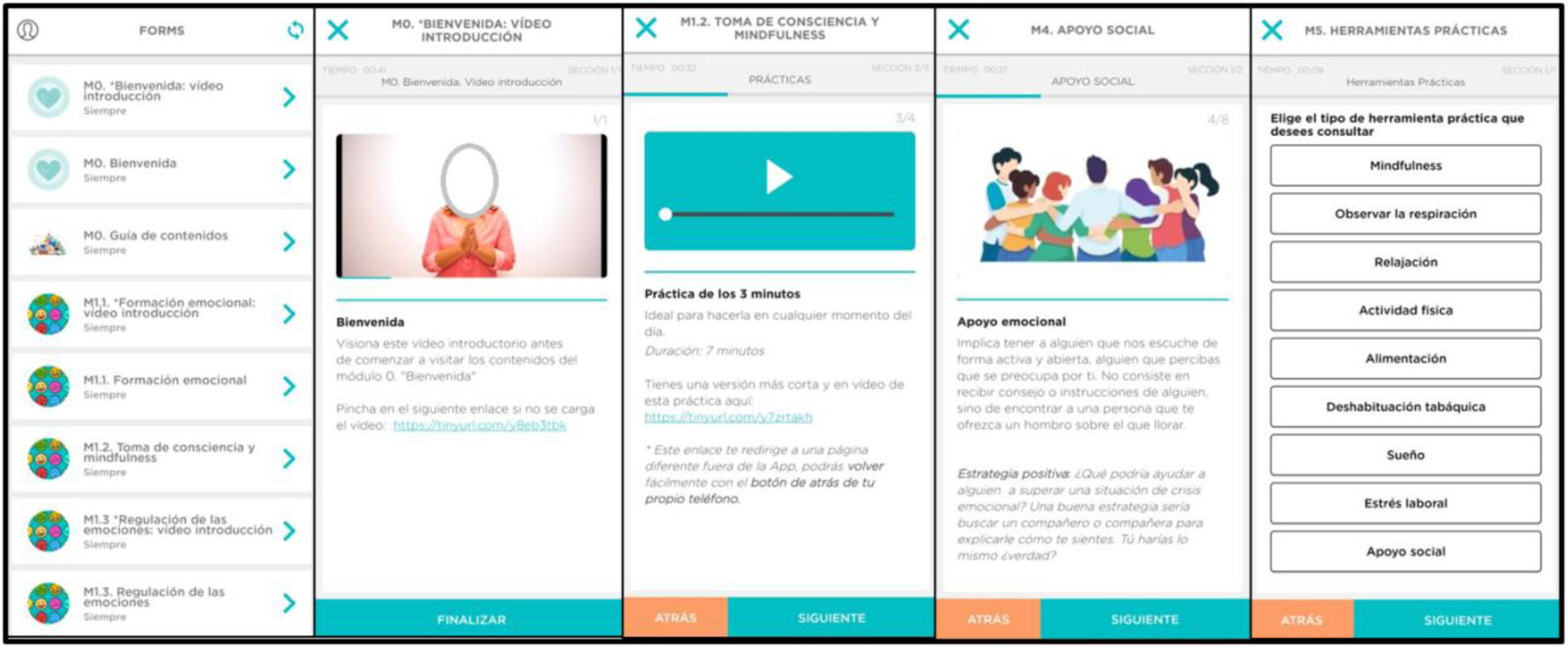
Visualization of permanent, generic (not-tailored) contents of the PsyCovidApp intervention.

Additionally, the intervention includes 14 temporal modules, which are available only for 24 hours. Each day the users will be prompted by a notification indicating that a new message is available. These messages contain a brief question followed by a short message. The messages have specific purposes, including: monitoring of mental health status, invitation to practice, reminders, and encouragement. The majority of the temporal modules offer tailored information or recommendations based on users’ responses to the brief questions (Figure 3).

**Figure 3.**
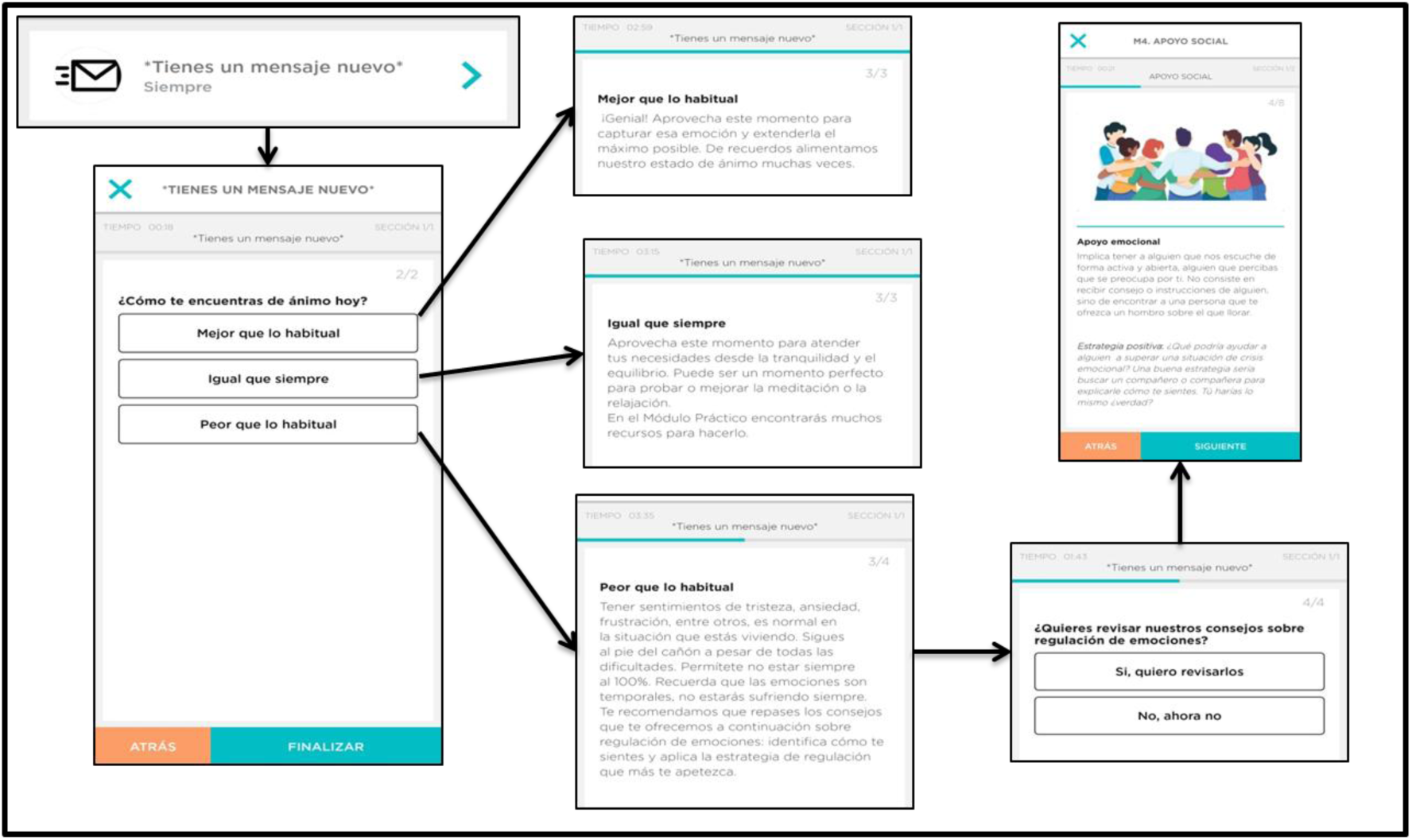
Visualization of temporal, tailored contents of the PsyCovidApp intervention.

#### Box 1.

**Description of the content of the PsyCovidApp intervention**

- Section 1. Emotional skills
- Knowing and identifying the most common emotional reactions that HCWs may experience during or after the COVID (depression, anxiety, acute and post-traumatic stress, and burnout)
- Introduction to mindfulness and audios to start its practice.
- Emotional regulation: strategies and practical advice (e.g., relaxation exercises through breathing or imagination, Jacobson’s progressive relaxation, etc.)
- Tips and tools to improve a period of crisis and/or emotional blunting.

Section 2. Lifestyle behavior

- Information on healthy lifestyle (i.e., physical activity, diet, exposure to sunlight, sleep and non-consumption of alcohol and tobacco) and its relationship with psychological well-being
- Self-assessment of a healthy lifestyle
- Tips to encourage support of healthy lifestyle behaviors.

Section 3. Work stress and burnout

- Informative content about work stress and burnout
- Practical advice to learn how to handle and prevent work stress and burnout.

Section 4. Social support

- Web resources to deepen the concept of social support and its different types
- Tips to promote social support and integrate it into the own code of social behavior.

Section 5. Practical tools

- Compilation of all the practical tools presented in the previous modules

### Description of the control App intervention

Participants in the control group will have access through the Clinicovery© App to a control App intervention (see Figure 4). This intervention will only include brief written information, adapted from a set of materials developed by the Spanish Society of Psychiatry for mental healthcare of HCWs during the COVID-19 pandemic. The information is organized in three sections: challenges faced by HCWs during the COVID-19 pandemic; common reactions to intense stress situations, and; mental health self-management recommendations. No temporal modules will be available for the control intervention.

**Figure 4.**
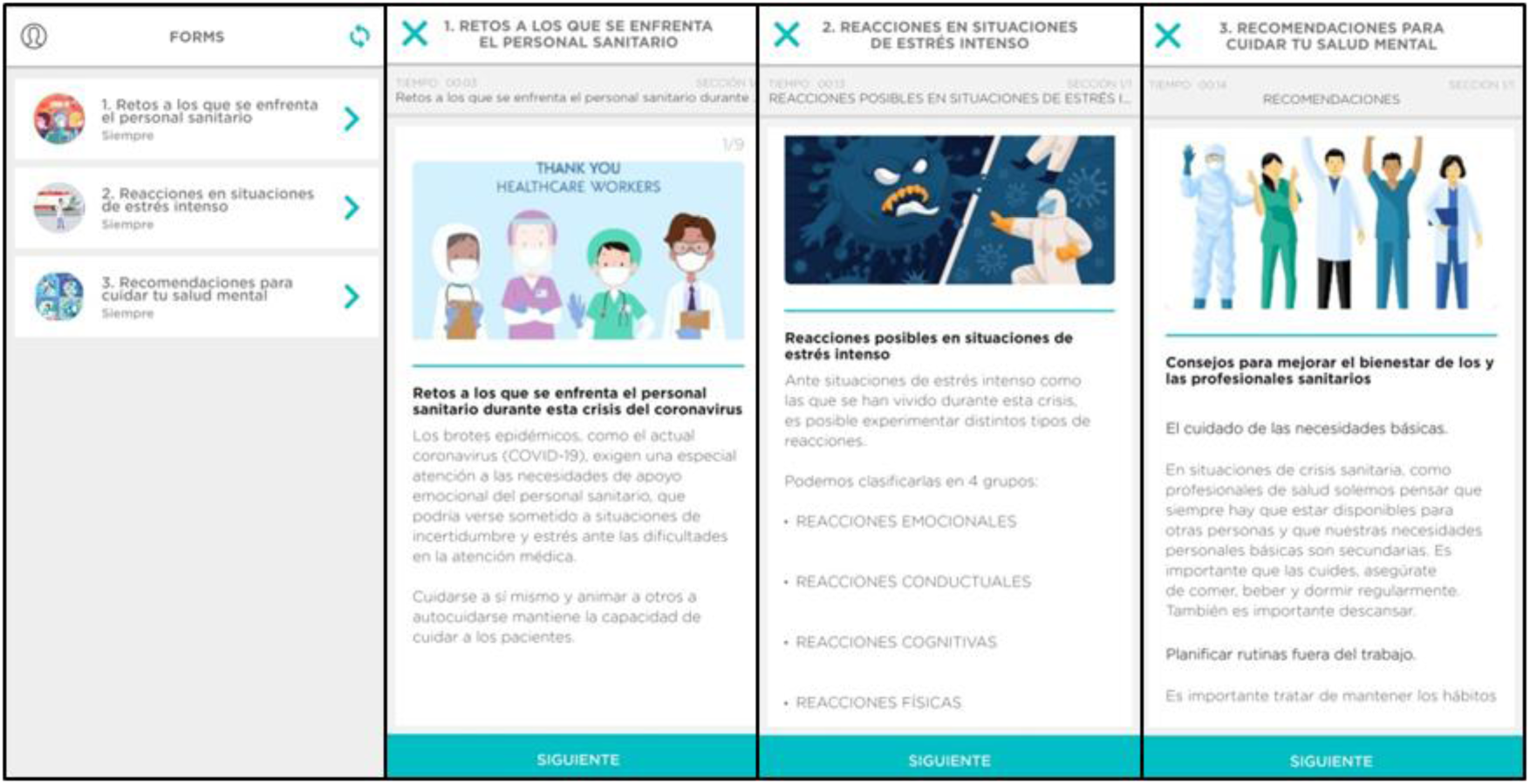
Visualization of the control intervention.

### Outcome measures

The primary outcome will be the difference between the intervention and control groups in the mean overall score the Depression, Anxiety and Stress Scales (DASS21) instrument^26^. The score ranges from 0 (worst outcome) to 21 (best outcome). The instrument contains three 7-items scales, assessing presence and intensity of depression, anxiety and stress. Items are based on a Likert-scale ranging from 0 – 3 points.

Secondary outcome measures will be the difference between intervention and control groups in the mean scores of the following instruments:

- Davidson Trauma Scale (DTS) ^27^. The DTS is a 17-item, Likert-scale, self-report instrument that assesses the 17 DSM-IV symptoms of post-traumatic stress disorder. Both a frequency and a severity score can be determined. The DTS yields a frequency score (ranging from 0 to 68), severity score (ranging from 0 to 68), and total score (ranging from 0 to 136). Higher scores are indicative of a worse outcome. The scope of the questionnaire to capture only post-traumatic stress disorders related with the COVID-19 health emergency.
- Maslach Burnout Inventory - Human Services Survey (MBI-HSS) ^28^. The MBI-HSS is a 22-item, Likert-scale, self-reported instrument that assesses three domains of burnout: emotional exhaustion (9 items), depersonalization (5 items) and personal achievement (8 items). All MBI items are scored using a 7-level frequency scale from “never” to “daily.” Each scale measures its own unique dimension of burnout. Scales cannot be combined to form a single burnout scale.
- Insomnia Severity Index (ISI)^29^. The ISI is a 7-item, Likert-scale, self-reported instrument assessing the severity of both night-time and daytime components of insomnia. Scores range from 0 (best outcome) to 28 (worst outcome).
- General Self-Efficacy Scale (GSE)^30^. The GSE is a 10-item, Likert-scale, self-reported instrument that assesses optimistic self-beliefs to cope with a variety of difficult demands in life. Scores range from 10 (worst outcome) to 40 (best outcome).
- System Usability Scale (SUS)^31^. The SUS is a 10-item, Likert-scale, self-reported instrument assessing subjective assessments of usability. It comprises three domains: effectiveness (whether users can successfully achieve their objectives); efficiency (how much effort and resource are expended in achieving those objectives); and satisfaction (whether the experience was satisfactory). Scores range from 0 (worst outcome) to 100 (best outcome).

### Sample size

It is estimated that 440 participants (220 per group, allowing for 10% attrition) will be required to detect at least an effect size of 0.25 (Cohen’s d) on DASS21 with 80% power and alfa of 5% (one-sided).

### Statistical analysis

The primary statistical analysis will be carried out on the basis of intention-to-treat (ITT) (i.e. all participants that agreed to participate will be included in the analysis according to the group to which they were assigned). The study results will be reported in accordance with the CONSORT 2010 statements^32^ and a full detailed statistical analysis plan will be prepared before recruitment starts (including any interim, subgroup and sensitivity analyses).Differences between groups of primary and secondary outcomes will be analysed using General Linear Modelling (ANCOVA) for continuous variable, adjusted by baseline score. The results from the trial will be presented as regression coefficient for predicting change in primary and secondary outcomes with 95% confidence intervals. We will use multiple imputation by chained equations (MICE) to fill in missing values (50 imputation sets)^33^.We will carry out subgroup analyses to examine the impact of the PsyCovidApp intervention on primary and secondary outcomes according to groups of HCWs based on the following baseline characteristics: use of psychopharmaceutical drugs (yes vs. no), use of psychotherapy (yes vs. no), and symptomatology of depression, anxiety, stress (yes vs. no - based on baseline DASS-21 median score).

### Ethical considerations

Research ethical committee approval was obtained by the Research Ethics Committee of the Balearic Islands (CEI-IB Ref No: IB 4216/20 PI). All potential participants submitting their expression of interest to participate in the study will receive a Participant Information Sheet. An audio-recorded informed consent will be obtained from every participant before data collection via telephone. HCWs will be informed of freedom to withdraw at any time and will be assured of anonymity by using special code numbers to identify themselves. All of the collected data will be pseudo-anonymized and kept confidentially. Only members of the research team will be able to re-identify the participants.

### Validity and reliability

This study uses a rigorous research design, an RCT with a representative and predetermined sample. It uses instruments with a high validity and reliability, and statistically analysis, which can be seen to reduce bias effectively and enhance the generalizability of research results beyond the target population. Moreover, trial participants, outcome assessors and data analysts of the research will be blinded to intervention allocation to reduce the biases in the evaluation of the effects of the intervention. The study design, procedures and reporting will follow the CONSORT statement recommendations on randomized controlled trials^32^.An additional strength of this study is that it will be performed under routine clinical conditions, and with a broad range of HCWs. This feature will give strong external validity.

## DISCUSSION

The global health emergency generated by the COVID-19 pandemic is posing an unprecedented challenge to frontline HCWs, who are facing high levels of workload under psychologically difficult situations with scarce resources and support.

There is a growing interest in the use of digital technology to deliver mental health care. However, this area of research is still in progress, and rigorous trials are needed to determine the extent to which these interventions can produce the desired benefits. This study will evaluate a psychoeducational mental health app based on CBT and mindfulness approaches, specifically developed to meet the needs of HCWs during the COVID-19 pandemic. This study will help to increase scientific evidence regarding the effectiveness of this type of intervention on this specific population and context. MHealth interventions are already being widely implemented because they are low cost, sustainable and highly scalable, but in absence of solid evidence about its effectiveness. The findings from this study will help health services and organizations to make informed decisions in relation to further development and roll out of this type of interventions, allowing them to ponder not only their attractive implementability features, but also providing robust data on impact on mental health.

### Limitations

The study has also some limitations. First, the two weeks follow-up period may be not enough to detect clinically meaningful differences in the selected outcomes. Although adherence to mHealth apps generally decrease overtime^34^, and two weeks is enough to access to all the contents of the PsyCovidApp intervention, a longer period of time may be needed to produce the desired positive effects on mental health. Second, restricting the study to HCWs with a smartphone and able to download and use mobile Apps may cause a selection bias which could reduce the generalizability of our results. This is a common limitation of mHealth trials, and which is unlikely to significantly affect the results of our study, since Spain is currently the country with higher smartphone use rates in the world, with 98% of users^20^.Third, the mental health of the participants will not be evaluated through a clinical interview, but rather using instruments indicated for symptomatology assessment rather than for clinical diagnosis. Fourth, we will not restrict our sample to HCWs with mental health problems at baseline. Including a large proportion of participants with no (or minor) mental health problems in our study may limit our ability to observe mental health improvements. Fifth, high dropout rates and low intervention adherence are common limitations of trials of low-intensity interventions, such as the one proposed in our study. Sixth, our ability to conduct the planned subgroup analyses may be limited by the size of such groups. Seventh, it is worth noting that this trial will be undertaken during a very specific and rapidly evolving context: the COVID-19 pandemic. Therefore, rapid recruitment of HCWs will be needed to ensure the intervention is homogeneously tested in the same context. Although we will allocate resources to recruit through hospitals, professional and scientific societies and HCWs unions, the feasibility of such rapid recruitment has not been previously examined. Finally, we will not be able to monitor the level of use of the App during the trial, and therefore it will not be possible to determine the extent to which higher intervention adherence is associated with higher benefits on mental health.

## CONCLUSION

This research will study for the first time the impact of a psychoeducational CBT-and mindful-based mHealth intervention specifically designed to protect mental health of frontline HCWs fighting against the COVID-19 pandemic in Spain. The findings from this study will be used to inform decisions about wider rollout of the PsyCovidApp intervention immediately after the trial. In addition, the study findings will help increase the scientific evidence concerning the impact of mental mHealth interventions on a specific population (HCWs) under a specific context (the health emergency caused by the COVID-19 pandemic); as well as, more generally, the evidence about the effectiveness of mHealth– an area of research still in its early stages, for which robust trials are urgently needed.

## Data Availability

Not applicable - study protocol, not involving collection of primary data at the protocol stage

## ACKNOWLEDGMENTS

We thank the HCWs who participated in the individual qualitative interviews to inform the design of the PsyCovidApp intervention.

## AUTHOR CONTRIBUTIONS

Study design: IRC, MJS, MAF, JGC. Manuscript preparation: IRC, MJS, IRP, MAF, CS, EGB. Manuscript revision and approval: all authors.

